# Healthcare Stakeholder Perspectives on a Value Assessment Approach for Duchenne Muscular Dystrophy Therapies

**DOI:** 10.1101/2023.07.11.23292507

**Authors:** Ryan Fischer, Pat Furlong, Annie Kennedy, Kelly Maynard, Marissa Penrod, Debra Miller, Chamindra G. Laverty, Linda Lowes, Nancy L. Kuntz, Perry B. Shieh, Jane Kondejewski, Peter J. Neumann, Jason Shafrin, Richard J. Willke

## Abstract

**BACKGROUND:** Traditional value assessment frameworks are challenged in comprehensively assessing the societal value new therapies bring to individuals with rare, progressive, genetic, fatal, neuromuscular diseases such as Duchenne muscular dystrophy (DMD).

**OBJECTIVE:** To identify how value assessment frameworks may need to be adapted to measure the value to society of DMD therapies.

**METHODS:** Three groups of stakeholders (patient advocates, clinicians, health economists) participated in semi-structured interviews around the International Society for Pharmacoeconomics and Outcomes Research’s Value Flower, which includes elements of value that can be considered within value assessments of healthcare technologies.

**RESULTS:** All stakeholders agreed that traditional value assessment frameworks based on the quality-adjusted life year (QALY) are narrow and will undervalue new DMD therapies. All stakeholders expressed some level of concern that use of the QALY as a key metric of value discriminates against patients with severe progressive diseases and disabilities. Some stakeholders saw value in using the QALY for cross-disease comparisons in resource-constrained environments if the methodology was appropriate. All stakeholders recommended considering additional elements of value in decision-making around new DMD therapies. These elements reflect: the economic and humanistic costs incurred by patients, caregivers, and families with Duchenne, such as indirect out-of-pocket costs, lost productivity, and family spillovers; the attributes that are meaningful for individuals with disabilities and high unmet need, such as severity of disease, value of hope, and real option value; and factors that contribute to improvements in population health, such as insurance value, equity, and scientific spillovers.

**CONCLUSIONS:** These findings highlight the need to expand traditional value assessment frameworks and take a holistic approach that incorporates the perspectives of individuals with Duchenne, caregivers, clinicians, and heath economists when assessing the societal value of new DMD therapies. Broadening value assessment will prevent restricted or delayed access to therapies for individuals with Duchenne.

## INTRODUCTION

Duchenne muscular dystrophy (DMD) is an X-linked disease caused by mutations in the *DMD* (dystrophin) gene. This progressive, neuromuscular disease is typically diagnosed at 4-5 years of age and impacts an estimated 1 in 3,500-6,000 male births [1, 2]. Affected individuals show deteriorating motor function with decline in ambulatory function during childhood [1, 3]. Upper limb weakness also develops, along with cardiac and respiratory complications, and these issues contribute to mortality by early adulthood [1, 4]. There is no cure for DMD, but treatments such as cardiac medications, steroids, and physical/occupational therapy are used to manage disease progression [5, 6]. Additional treatment options, including exon-skipping therapies, stop codon readthrough medications and gene therapy, are emerging [7-9].

The continual development and advancement of new healthcare technologies has led to the creation of frameworks to measure value in health [10]. Traditional value assessment frameworks have primarily focused on net costs and measured health benefits to patients. Health benefits to patients are often quantified using quality-adjusted life years (QALYs). QALYs are a measure of survival time weighted by the quality of life during that time; the quality of life weighting goes from 0 (death, or the worst imaginable health state) to 1 (full healthy life). Quality of life can be measured in a variety of ways but is most commonly assessed with the EuroQol-5Dimension (EQ-5D). The EQ-5D is a disease agnostic instrument that enables comparisons across a wide range of disease areas; however, it may be insensitive to particular aspects of certain diseases [11]. One QALY can be 1 year of perfect health, or 2 years at a 0.5 quality of life rating, or an equivalent combination. The ratio of change in costs (related to treatment and its outcomes) to the change in QALYs due to treatment (i.e., cost/QALY metric) is known as an incremental cost-utility ratio and is often used as a “starting point” to inform value decisions [12]; however, QALYs may not comprehensively capture all potential benefits to patients, payers, families, or society [13].

Thought leaders have proposed broadening and advancing the view of value in health beyond the QALY to assess the societal value of new healthcare technologies. In 2016, the Second Panel on Cost Effectiveness in Health and Medicine provided updated guidance on the evolution of cost-effectiveness analysis (CEA), recommending two reference case analyses, one from a healthcare sector perspective and another from a societal perspective, and the use of an “impact inventory” as a checklist of a healthcare technology’s health and non-health impacts [14]. In 2018, a special task force of the International Society for Pharmacoeconomics and Outcomes Research (ISPOR) created the ISPOR Value Flower, which includes 12 elements of value (petals) that can be considered within value assessments of healthcare technologies. The ISPOR Value Flower and its recent adaptations (**Figure S1**) highlight common but inconsistently used elements, as well as novel elements of value that have previously been underappreciated by many payers (**Table S1**) [15-17]. Including novel elements of value in value assessment frameworks may have particular relevance for individuals with rare diseases and transformative healthcare technologies such as cell and gene therapies [16].

This manuscript reports the elicited perspectives of three groups of stakeholders (patient advocates, clinicians, and health economists) on the inclusion of specific elements of value from the ISPOR Value Flower in a value assessment framework for DMD therapies. These perspectives should guide decision-makers to ensure they have incorporated all perspectives and elements of value in their value assessment frameworks for new DMD therapies. Findings should promote further debate and comprehensive evidence generation to establish an evolved and holistic approach to valuing new DMD therapies.

## METHODS AND MATERIALS

This initiative evaluated the importance of the elements of value from the ISPOR Value Flower [15,17] for assessing new DMD therapies through semi-structured interviews with patient advocates, clinicians, and health economists. The study was conducted in four phases: 1) selection, 2) preparation, 3) interviews, and 4) review and clarification. Each phase took approximately one month to complete. The study methodology and phase objectives are provided in **Figure 1**.

**Figure 1:**
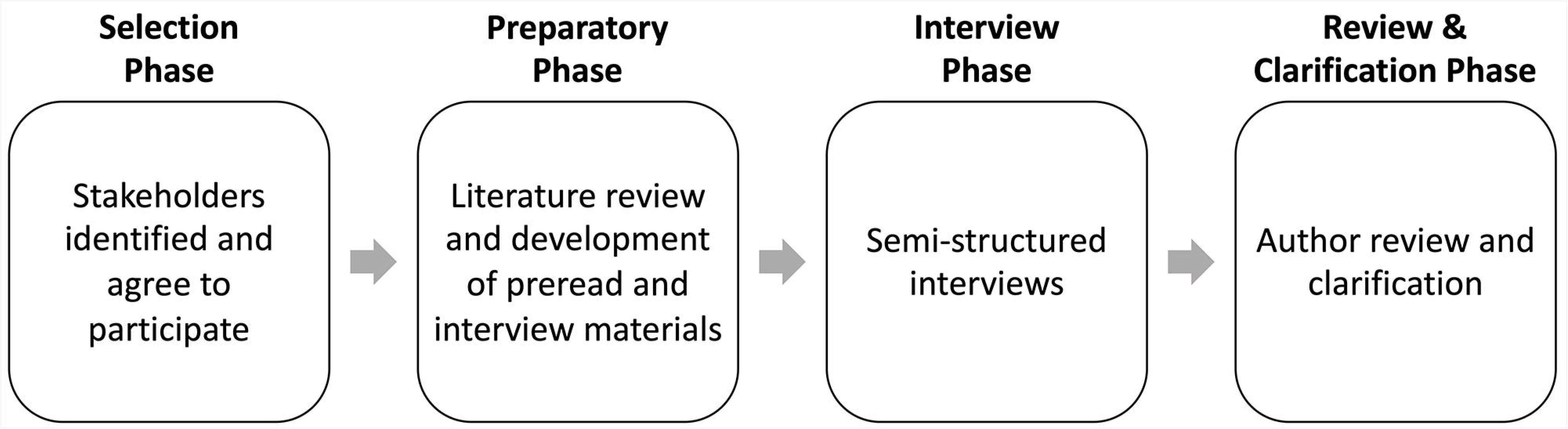
Methodology.

### The Selection Phase

The selection phase identified stakeholders in three areas: patient advocacy, clinical, and health-economics, who were willing to share their perspectives. These diverse groups were intentionally engaged to provide a scientific, balanced approach and reduce any bias. Stakeholders were selected using the following criteria. Patient advocates were considered if they were involved with or had experience working for a DMD not-for-profit organization, exposing them to a broad cross-section of the DMD patient and caregiver community. Clinicians were considered if they had extensive experience diagnosing and treating individuals with Duchenne. Health economists were considered if they had expertise associated with value assessment frameworks, including the ISPOR Value Flower, and understood the applicability of these frameworks for gene therapies; had expertise in value assessment frameworks for therapies for chronic diseases, which allowed them to provide informed insight on expanded value assessment frameworks for rare, severe progressive diseases, including DMD; and had knowledge of how value assessment frameworks can influence reimbursement decisions. In total, 13 stakeholders were included in the study: 6 patient advocates, 4 clinicians, and 3 health economists.

### The Preparatory Phase

The preparatory phase consisted of a targeted literature review focused on articles published after the development of the ISPOR Value Flower. The search strategy included the keywords: value framework, rare disease, elements of value, Duchenne muscular dystrophy, health technology assessments. The review identified 186 relevant journal articles. These articles were used to create a 20-page preread. The preread provided an overview of the initiative and a description of each element of value from the original and updated ISPOR Value Flower [15-17] and their relevance to DMD. Stakeholders had 2 weeks to review the preread prior to their interviews.

### The Interview Phase

Interviews were semi-structured, conducted remotely, recorded for accuracy and data collection, and facilitated by the same lead interviewer and medical writer. Interviews were guided by slides that followed a format similar to the preread. The interviewer started each interview by providing an overview of the ISPOR Value Flower. Three questions were presented to all stakeholders on each element of value to facilitate discussion: ‘*Should this element of value be included in a value assessment framework for DMD therapies? How does inclusion of this element of value impact you and your peers? What are the barriers to uptake of this element of value?’* Stakeholders were asked to prioritize elements of value relevant to a value assessment framework for DMD therapies, from their perspective. Deviations to the slide order were permitted and determined by stakeholder interests or questions. The interviews were conducted between March 1, 2023, and March 22, 2023.

### Review and Clarification Phase

Findings were summarized by element of value from the ISPOR Value Flower and aggregated by stakeholder. Summarized findings were reviewed by the stakeholders, who were provided the opportunity to clarify statements and were offered additional time to ask questions or provide more context or information on statements made during the interviews. Stakeholders provided minor feedback, and one health economist requested another meeting. All stakeholders (authors) retained control over the content of this publication.

This study represents stakeholder opinions and did not require Institutional Review Board approval.

## RESULTS

The main perspectives that emerged from the stakeholder interviews are described below (**Figure 2**), and examples of quotes are provided in **Tables 1-3**.

**Figure 2:**
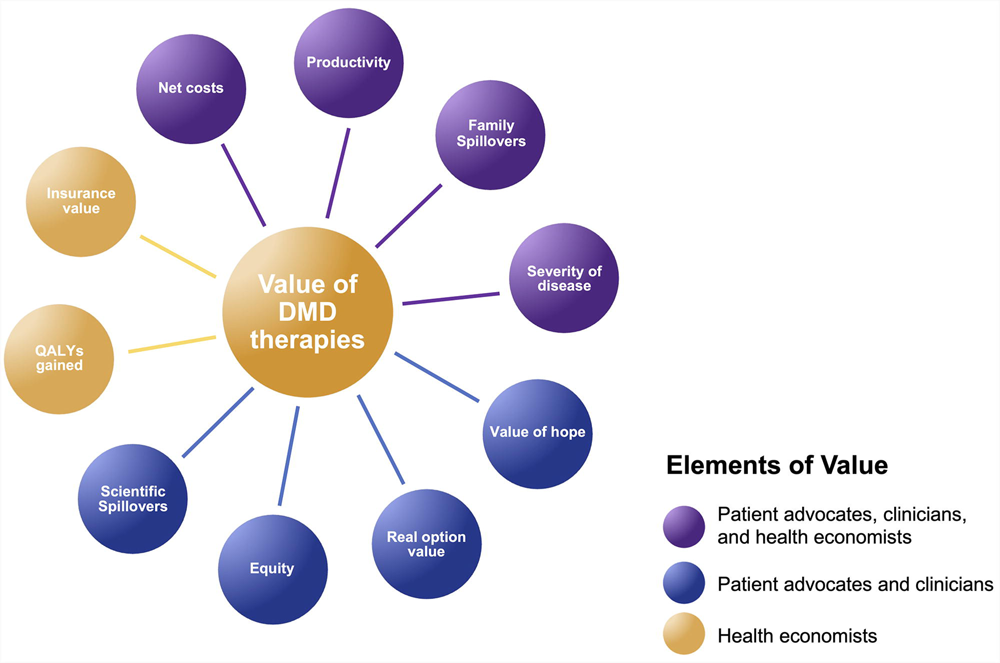
Proposed Value Assessment Framework for DMD Therapies: Adapted from the ISPOR Value Flower [15-17]. DMD, Duchenne Muscular Dystrophy; ISPOR, International Society for Pharmacoeconomics and Outcomes Research; QALY, quality-adjusted life year.

**Table 1:**
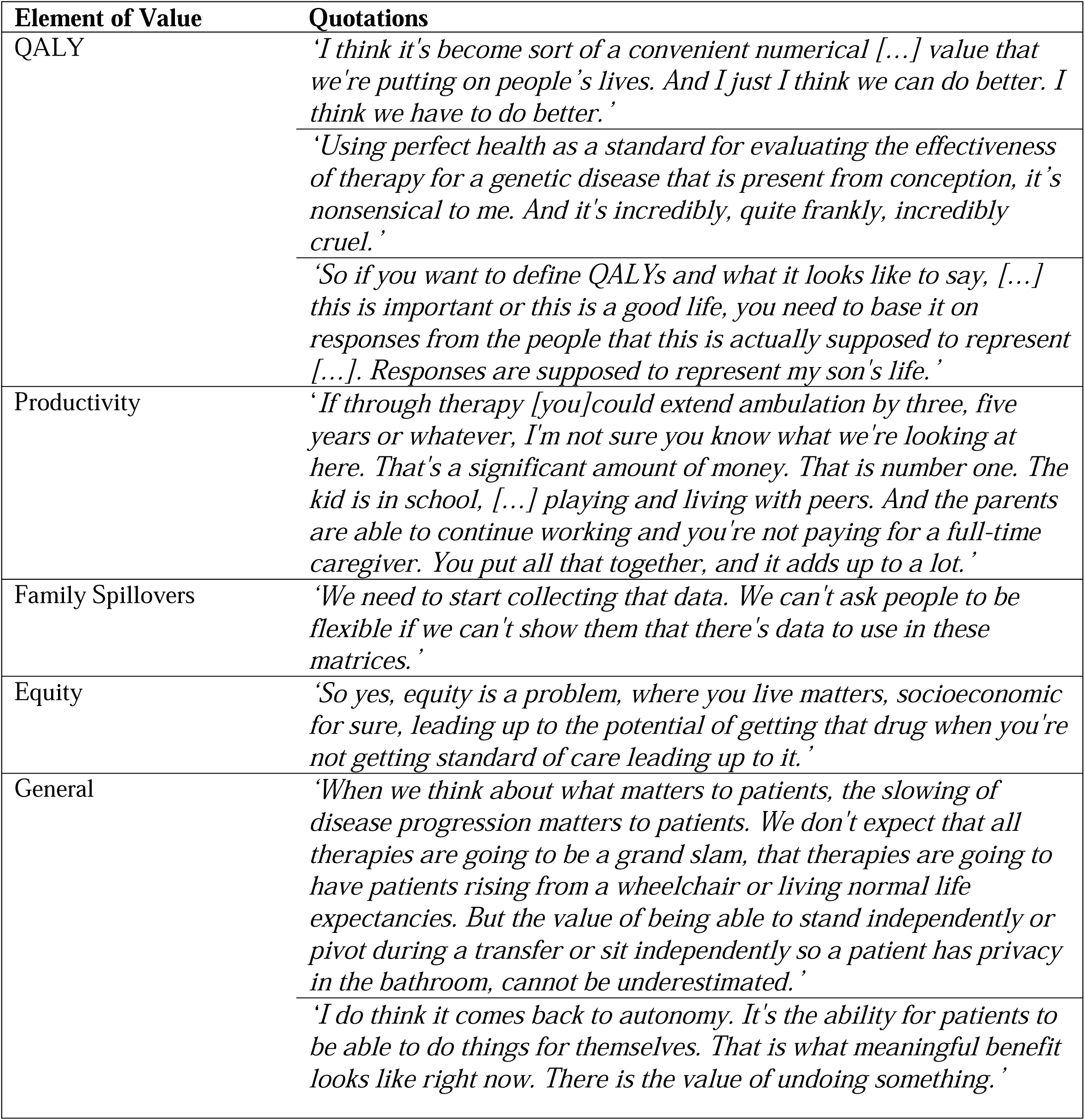
Quotes by element of value from the patient advocate perspective.

| Element of Value | Quotations |
| --- | --- |
| QALY | *'I think it's become sort of a convenient numerical [...] value that we're putting on people's lives. And I just I think we can do better. I think we have to do better.'*<br>*'Using perfect health as a standard for evaluating the effectiveness of therapy for a genetic disease that is present from conception, it's nonsensical to me. And it's incredibly, quite frankly, incredibly cruel.'*<br>*'So if you want to define QALYs and what it looks like to say, [...] this is important or this is a good life, you need to base it on responses from the people that this is actually supposed to represent [...]. Responses are supposed to represent my son's life.'* |
| Productivity | *'If through therapy [you] could extend ambulation by three, five years or whatever, I'm not sure you know what we're looking at here. That's a significant amount of money. That is number one. The kid is in school, [...] playing and living with peers. And the parents are able to continue working and you're not paying for a full-time caregiver. You put all that together, and it adds up to a lot.'* |
| Family Spillovers | *'We need to start collecting that data. We can't ask people to be flexible if we can't show them that there's data to use in these matrices.'* |
| Equity | *'So yes, equity is a problem, where you live matters, socioeconomic for sure, leading up to the potential of getting that drug when you're not getting standard of care leading up to it.'* |
| General | *'When we think about what matters to patients, the slowing of disease progression matters to patients. We don't expect that all therapies are going to be a grand slam, that therapies are going to have patients rising from a wheelchair or living normal life expectancies. But the value of being able to stand independently or pivot during a transfer or sit independently so a patient has privacy in the bathroom, cannot be underestimated.'*<br>*'I do think it comes back to autonomy. It's the ability for patients to be able to do things for themselves. That is what meaningful benefit looks like right now. There is the value of undoing something.'* |

**Table 2:** Quotes by element of value from the clinician perspective.

| Element of Value | Quotations |
| --- | --- |
| QALY | *‘If you’re talking about value, it’s the first thing that comes to people’s mind, QALYs. It’s hard to ignore QALYs because everyone talks about QALYs and it’s kind of the language that everyone speaks.’* |
|  | *‘You ask him, are you able to do the things you enjoy? They say absolutely. Because what they enjoy is video games. They might have enjoyed soccer more. But there’s no way for them to know that.’* |
| Severity of Disease | *‘I would almost encourage the severity of disease category to be broadened to include special attention to progressive disorders - those disorders that are associated with active progression or deterioration of tissue and function.’* |
|  | *‘They didn’t have poor behavior or poor health or poor lifestyle; this was just given to them.’* |
| Scientific Spillovers | *‘I think that the potential scientific gain is immense. So I do think that there’s a lot that is to be gained from really tackling this complicated problem.’* |

**Table 3:** Quotes by element of value from the health economist perspective.

| **Element of Value** | **Quotations** |
| --- | --- |
| QALY | *‘The QALY could do other things, and that’s where I think some of this conversation may need to go – how do you incorporate disease-specific utilities into this so that they do a better job of capturing the subtleties of disease relative to the five usual dimensions that often go into utility measure.’* |
| Net Costs | *‘You can certainly say we want to weigh all the costs and all the benefits. Certainly this can go into your caregiver costs or the home remodeling.’* |
| Productivity | *‘I think these arguments around productivity and all these other costs are very compelling. And the fact that payers don’t pay for it doesn’t mean they don’t have value.’* |
| Family Spillovers | *‘I think the spillover on kids and grandparents as well, if they are retired, but now they have to spend a lot of time caregiving. OK. That’s in theory measurable, but in practice is just difficult, not impossible, but it’s difficult. So even though those concerns are very real and probably are true, it’s just hard to measure them. But again if that’s possible, it certainly could be relevant.’* |
| Insurance Value | *‘It can be high, especially for rare diseases and things that are really severe because that’s typically what you want to insure against.’* |
| Severity of Disease | *‘The reason I put disease severity high is I think there’s empirical evidence to back some of these statements here that moving from a utility value of 0.3 to 0.4 is worth more to patients than moving from a value of 0.8 to 0.9. Therefore, we might need to modify our cost-effectiveness thresholds, for example, allowing higher thresholds for more severe diseases.’* |
| Value of Hope | *‘I think there are surveys to be done to really understand better preferences around risk taking and treatment effects.’* |
| Real Option Value | *‘It’s just usually there’s not good data on that. If keeping you at a good health state, [...] if it’s a progressive disease, that certainly would have option value. If there’s a new treatment.’* |
| Equity | *‘So I think the key thing is that the treatment helps reduce the disparity.’* |
| General | *‘I think you always start with the basics of functional improvement, and then you capture some of the key costs and just lay out the pieces that you’ve measured so they see the profile. And then you can put them together, if you want, into cost-effectiveness, but clearly the payers are going to be interested in the underlying facts.’* |

### QALYs

The QALY is the fraction of a perfectly healthy life-year that remains after accounting for the damaging effects of an illness or condition [15]. Some patient advocates suggested that the use of the QALY in a value assessment framework for DMD therapies should be banned. Others recognized that QALYs can inform healthcare resource allocation decisions. All patient advocates felt that the QALY has no relevance for individuals and families living with a rare, progressive, debilitating disease, and should not be used as a singular tool to value DMD therapies. They recommended a DMD-specific QALY based on disease-specific quality of life assessments, noting that the QALY does not account for transition and adjustment in DMD or allow for any nuance. The patient advocates stated that the QALY is not appropriate for a pediatric disease, where quality of life assessments may be based on parents speaking for their children, which creates an inherent bias.

The clinicians acknowledged the need for a metric when assessing the value of DMD therapies, but agreed that the QALY is imperfect. An individual’s quality of life is not very well captured in a QALY, especially if they have a chronic progressive disease. The clinicians recommended that quality of life assessments in DMD could include questions related to education, employment goals, aspirations, pain, and happiness. The assessments should be flexible and able to capture the impact of extending life in DMD, especially as new standards of care, such as synthetic corticosteroids, and new therapies with additive value will continue to change the natural history of disease. The clinicians stated that quality of life in DMD should not be captured by disease-agnostic instruments that include insights from the general population, because perceptions of good quality of life differ depending on an individual’s experiences and circumstances; for example, the general population will rate life in a wheelchair much more negatively than an individual with Duchenne.

The health economists mentioned that the use of the QALY to value healthcare technologies has been debated and controversial for many decades; however, the QALY is just one input used in health economic analyses and it is never used rigidly by payers to make judgments. The health economists noted that the QALY is used to value healthcare technologies, not individuals. The QALY is flexible and able to capture various aspects of different diseases, including heterogeneity, if the methodology is appropriate. Disease-agnostic instruments—such as the EQ-5D—to measure quality of life impacts can be used across diseases; however, these disease-agnostic instruments may not be sensitive enough to capture the subtleties of many diseases, including DMD. This may be addressed by the collection of quality of life estimates from disease-specific instruments; collecting quality of life data, however, can be difficult in pediatric rare diseases due to small sample sizes and the challenging nature of eliciting utilities from children.

### Net Costs

Net costs are the intervention costs minus averted medical and productivity costs [15]. All stakeholders recognized the importance of net costs in a value assessment framework for DMD therapies. In DMD, net costs should include the out-of-pocket costs beyond drug costs that are sustained by individuals with Duchenne and their families, such as traveling to doctors’ appointments, out-of-pocket medical expenditures, copays for medical services, the cost of wheelchairs, wheelchair accessible vans, other equipment, home adaptations or relocating homes, and the cost of extraordinary disability, including the requirement for a reliable and knowledgeable caregiver to be with an individual with Duchenne 24 hours a day. In addition, there are the costs of school support financed by state programs. The patient advocates stated that commercial payers and federal and state programs want to understand the return on their investment, including benefits, such as improvements in patients’ health, and spending from Medicaid, the school system, or waivers. All stakeholders agreed there is a need for more research capturing out-of-pocket costs beyond drug costs in DMD, which are mostly underappreciated by payers. The National Economic Burden of Rare Disease Study, which evaluated direct, indirect, and non-medical costs of rare disease in the US in 2019 and included individuals with Duchenne, estimated that nearly 60% of costs were associated with expenses shouldered by families or society [18].

### Productivity

Productivity is the measurement and value of productivity gains and losses due to healthcare interventions [15]. All stakeholders indicated that work productivity for individuals with Duchenne and their caregivers should be included in a value assessment framework for DMD therapies. This productivity is likely to increase as new therapies extend quantity and quality of life. Individuals with Duchenne can attend college and contribute to society even if they are physically impaired, and society has a responsibility to enable their productivity by reducing stigma and providing reasonable support. A value assessment framework must capture the nuances of DMD, including the value of an individual with Duchenne retaining their arm strength and the independence that allows them to go to college while their parents can continue to work. The clinicians recommended collecting data using measures of productivity. Health economists suggested that modeling the impact of new DMD therapies on long-term productivity and lost opportunity may involve a lot of assumptions and may not be considered reliable by payers. They noted that productivity costs may be excluded from value assessment frameworks by payers if they have not been measured in a robust manner. The National Economic Burden of Rare Disease Study used on online survey to estimate cost due to reduced labor market participation, productivity loss for those in the labor force, non-medical costs of rare diseases (such as the cost of hiring professional non-medical caregivers to assist with daily living, necessary home modification costs), and disability benefits [18].

### Family Spillovers

Family spillovers are the impacts of illness extending beyond the patient to unpaid caregivers and other family members who commonly incur out-of-pocket and time costs, lost productivity, and decreased health-related quality of life caring for and caring about a sick family member [19]. All stakeholders acknowledged that family spillovers are highly relevant to determining the value of DMD therapies, noting that earlier intervention and improved outcomes may reduce the impact of DMD on families. Stakeholders indicated that DMD affects every family member’s productivity, employment, education, and physical and mental health. Parents of individuals with Duchenne may be forced to neglect careers and experience financial difficulties. Siblings of individuals with Duchenne may lose time with friends and often give up activities. Families frequently rely on support from grandparents or social contacts, if available. Family life is disrupted as there is less ability to plan. Families must change their behaviors when individuals with Duchenne transition to adulthood. There may be parent-child conflict as individuals with Duchenne desire more independence but may experience social isolation due to their limited mobility and need for more physical support. The clinicians mentioned that the rate of divorce may be higher in families with Duchenne compared to other genetic conditions with a recessive inheritance. In DMD, an X-linked recessive disease, the mother is the carrier in approximately 65% of cases [20], could hold guilt and be or feel blamed, and may have manifesting health issues herself. The clinicians recommended measuring the impact of DMD on parents and caregivers with a musculoskeletal pain score, emotional pain score, or a depression scale. The health economists stated that family spillovers are measurable in theory, but this may be difficult in practice. It may be possible to model treatment effects to reduce impacts on the family. One approach would be to show an association between severity of disease and negative impacts on the family, and that treatment slows progression.

### Severity of Disease

Severity of disease refers to the greater weight (for themselves) individuals place on improvements in health from more severe health states than on equivalent improvements from less severe states [15]. All stakeholders agreed that severity of disease contributes to the value of DMD therapies. DMD should be given special consideration as a progressive inherited disease where there is deterioration requiring extensive support for all activities of daily living, including eating, showering, and dressing; a shortened lifespan; and no curative treatment. Young boys with Duchenne can have difficulty keeping up with peers, loss of ambulation occurs around 10-12 years or age, and self-care such as feeding can become difficult in the teens to early twenties. The health economists indicated there may be a need to modify cost-effectiveness thresholds; for example, allowing higher thresholds for more severe diseases. New approaches such as disease severity modifiers—as adopted by the National Institute for Health and Care Excellence (NICE)—or novel approaches such as the Generalized Risk-Adjusted Cost-Effectiveness (GRACE) analysis — may be useful.

### Value of Hope

Value of hope refers to the general population-and /or patient-perceived trade-offs for a chance of cure or extended survival [15]. The patient advocates and clinicians noted that value of hope is relevant to a value assessment framework for DMD therapies. They emphasized that individuals and families with Duchenne have tolerance for uncertainty because the condition is fatal, and tolerance for uncertainty increases with disease progression. The patient advocates suggested that the value framework should label this element as ‘patient preferences’. They recommended that individuals and families with Duchenne be educated about newer therapies, since there may be a lack of understanding about the terminology around adverse events, and not enough knowledge of effectiveness when making choices among different therapies. The health economists indicated that value of hope usually applies when treatments are seen as potentially truly curative for a subset of treated patients, but this concept may also have relevance if new DMD therapies allow for the maintenance of functional autonomy in some subset of treated individuals with Duchenne. Value of hope may be particularly relevant when payers are considering new treatments with uncertain or highly variable outcomes across patients.

### Real Option Value

Real option value is generated when a healthcare technology that extends life creates opportunities for the patient to benefit from other future advances in medicine [15]. The patient advocates and clinicians recognized that real option value is important for determining the value of DMD therapies. Preserving function for an additional 6 to 12 months may enable an individual with Duchenne to receive and benefit from future therapy. Science is advancing, and current therapies buy time, especially if treatment is early enough to substantially slow disease progression. Drugs may combine, be additive and/or synergistic in terms of mechanism of action and ultimate impact. The nuances of slowing disease progression and maintaining independence are hard to capture but must be articulated to payers, as they can substantially reduce the impact and costs of DMD.

The health economists suggested that while real option value is highly relevant to DMD, quantifying this value in practice is challenging. Option value at time of drug launch is very conjectural and there are no good data on the effectiveness of future innovations. One could assume that future innovation improves health as much as that in the past, but this is also a conjectural assumption. While in most cases real option value produces positive value, it can also have negative aspects if a patient takes a treatment that precludes them from receiving other treatments in the future.

### Equity

Equity refers to the value of healthcare technologies that address equity across populations, including healthy versus sick [15]. The patient advocates and clinicians mentioned that families with Duchenne and other rare diseases are truly disadvantaged. Zip code can impact diagnosis, and there is inequity in access to insurance, knowledgeable healthcare, families’ abilities to navigate healthcare, and lost opportunities for individuals with Duchenne, including reduced chance for marriage, the inability to drive, and limitations in performing self-care. The value of providing more options to individuals with Duchenne should be included in a value assessment framework for DMD therapies.

The health economists emphasized the need to define the meaning of disparities (e.g., disparities across income groups, education levels, race/ethnicity, or challenges in access to healthcare) when including equity in a value assessment framework. They noted that Medicaid or state benefit programs could cover a treatment for low-income patients with limited access to healthcare, or broader coverage could protect patients with standard insurance from a decrease in income due to additional expenses associated with a disease.

The health economists mentioned that equity can only be considered an element of value if (i) disease incidence occurs across different groups (e.g., income, racial, ethnic), and (ii) the healthcare technology helps reduce the disparity either by working better for certain groups or because the disease incidence is more common among disadvantaged individuals. For example, sickle cell disease disproportionately affects African Americans. Improved treatments for sickle cell disease would help reduce disparities at a societal level.

### Scientific Spillovers

Scientific spillovers are the value of the impact a new healthcare technology will have on the development of other healthcare technologies in the future [15]. The patient advocates and clinicians appreciated that there is value in the approval of new DMD therapies, as they can lead to increased competition, expansion of innovative therapies, and long-term return on upfront investment. The patient advocates stated that a new drug approval can impact the design of clinical trials and increases payers’ understanding of drug development in a disease area, such as around the use of surrogate endpoints. As new therapies are approved, the scientific community could be motivated and investors may develop improved technologies, which could help to drive down costs. The clinicians recognized the scientific benefit of dystrophin-directed translational science and clinical trials in DMD, as DMD is a complicated, progressive disease with many different mutations of the dystrophin gene. Basic science may contribute to the overall body of knowledge in DMD and other rare diseases. The health economists noted that scientific spillovers are relevant when an approved new treatment for a disease generates real-world evidence that could be of value for other treatments in development.

### Insurance Value

Insurance value is the value of physical and financial risk protection of a new healthcare technology, whereby a new technology reduces the physical risk of getting or staying sick, offering “physical risk protection,” and the “financial risk protection” of greater options for consumers due to medical care, expanding the possibility of insuring against illness [15]. The health economists stated that insurance value should be considered in a value assessment framework for DMD therapies. They clarified that insurance value can be high for rare and severely progressive diseases because individuals typically want to insure against the worst outcomes (i.e., life threatening or highly debilitating diseases). The health economists cautioned that the concept of insurance value may be too complex for some payers to understand, although there is a lot of heterogeneity among payers in the US.

The patient advocates did not recommend insurance value as an element of value for DMD therapies. They conflated insurance value with frustration over insurance denials, and the time taken to deal with these denials, especially as individuals with Duchenne get older. The clinicians stated that although DMD is a rare disease, the high spontaneous mutation rate precludes prenatal awareness in families, making it among the more frequently occurring rare diseases; therefore, there may be value in it being covered in an insurance package.

### Other Elements of Value

Fear of contagion and disease is most commonly associated with infectious diseases, and it assesses the benefits of a healthcare technology that extend beyond the treated patient [15]. Reduction in uncertainty is the value given to a healthcare technology that improves the certainty of outcomes or appropriate use of therapies [15]. Adherence-improving factors are the value of offering advantages over existing alternatives, such as simpler dosing schedules or alternate routes of administration [15]. Fear of contagion and disease, reduction in uncertainty, and adherence-improving factors did not resonate with any of the stakeholders.

## DISCUSSION

This initiative provides new insights into the elements of value that patient advocates, clinicians, and health economists perceive as relevant for a value assessment framework for DMD therapies. There was substantial overlap in perspectives, with all stakeholders recommending inclusion of net costs, productivity, family spillovers, and severity of disease as the elements most likely to impact the value of new DMD therapies. The patient advocates and clinicians noted the importance of value of hope, real option value, equity, and scientific spillovers, but cautioned they may be difficult to quantify. The health economists were the only stakeholders to highlight the relevance of QALYs gained and insurance value. New DMD therapies may have the potential to delay disease progression, enable individuals with Duchenne to maintain autonomy longer, and increase life expectancy. Even small-to-moderate reductions in the rate of disease progression could provide parents and families time with their children, preserve the autonomy and independence of individuals with Duchenne and improve their quality and quantity of life, and increase the functioning and wellbeing of caregivers and families, substantially impacting the DMD community and society. Accurate and comprehensive assessment of the value of new DMD therapies is essential to prevent restricted or delayed access for individuals with Duchenne.

Most stakeholders acknowledged the cost/QALY metric as a useful starting point for a value assessment framework for DMD therapies, but all stakeholders identified challenges with evaluations using QALYs and the cost/QALY metric. Specifically, extending the lives of individuals with disabilities or underlying health conditions gains fewer QALYs than extending the lives of more healthy individuals if quality of life is measured from the general population perspective rather than patient perspective [21-23]. For this reason, the Inflation Reduction Act has barred the Centers for Medicare and Medicaid Services from using QALYs in setting a maximum fair price for high-cost drugs used by Medicare [24]. Consequently, there is a need to value improvement in quality of life, irrespective of the starting point, including improvement relative to the counterfactual. Disease-specific questionnaires can be used to measure improvement in quality of life, and the QALY can be used to value improvement in quality of life, if there is agreement on how ‘improvement’ is defined. Considerations of the lived experience of individuals with Duchenne could help produce accurate information on QALYs gained due to DMD therapies, leading to more precise assessments of value. These considerations are reflected in a recently developed 30-item, 6-category (autonomy, daily activities, feelings and emotions, identity, physical aspects, social relationships) framework that captures quality of life in DMD from the perspective of boys and men with the disease [25].

All stakeholders agreed that new DMD therapies can provide value by reducing net costs, increasing the productivity of individuals with Duchenne and their caregivers, and mitigating family spillovers. A recent study using claims and electronic medical records data from the Decision Resources Group’s Real World Data Repository (2011–2020) reported the average annual cost of medical care (medical and pharmaceutical claims) for an individual with Duchenne in the United States, accounting for disease progression, was $71,451 (2020 USD) [26]. The National Economic Burden of Rare Disease Study estimated the overall annual economic burden of rare diseases in the United States in 2019 exceeded $997 billion. Of the total economic burden, the largest costs were indirect costs from productivity losses at $437 billion, direct medical costs at $449 billion, and non-medical and uncovered healthcare costs of $111 billion absorbed directly by families living with rare diseases [18]. In DMD, indirect costs and productivity loss are highest when ambulation is lost. Families must purchase wheelchairs, make house adaptations, purchase or modify cars or vans [27], and provide full-time formal caregivers for older individuals with Duchenne. Caregivers often have to reduce their working hours or stop work completely to care for an individual with Duchenne. Reductions in predicted annual earnings for female caregivers of individuals with Duchenne with 0–3 y and ≥4 y of ambulation loss have been estimated at $13,828 and $23,995 (2020 USD), respectively [28]. Data on family spillovers is limited, as they are not often captured in claims data and existing databases. The EveryLife Foundation and Lewin Group have some information on family spillovers [18], and other organizations have conducted family surveys. Patient-centered value elements that could be incorporated in a value assessment framework for DMD therapies include the National Health Council’s Patient-Centered Core Impact Sets [29].

All stakeholders stated that severity of disease impacts the value of DMD therapies. Some health technology assessment agencies have begun incorporating a “severity premium” in their decision-making. The health economists suggested there should be a higher willingness to pay for treatments for more severe disease, citing NICE as an example. NICE is introducing a severity modifier into its methods and processes for health technology assessment, placing increased value on QALY gains in severe disease. For conditions considered more severe, QALY gains will be upweighted by a multiplier of 1.2 or 1.7. Eligibility for the multiplier will be determined by lifetime QALY shortfall, absolute or proportional, whichever implies greater severity level for the eligible patient population compared to the general population [30]. The QALY weight is multiplied by 1.2 if proportional QALY shortfall is 0.85-0.95 or absolute QALY shortfall is 12-18. The QALY weight is multiplied by 1.7 if proportional QALY shortfall is at least 0.95 or absolute QALY shortfall is at least 18 [30]. The health economists recommended the use of absolute QALY shortfall for a pediatric population.

Patient advocates and clinicians recognized value of hope, real option value, equity, and scientific spillovers as highly important for determining the value of DMD therapies. Patient advocates and clinicians agreed that individuals and families with Duchenne live with uncertainty, which increases with disease progression [31-34]. Individuals with Duchenne and caregivers will take the potential opportunity for gains in strength and cardiac and pulmonary function over other risks and uncertainties associated with treatments [32]. Health economists called for additional data collection around preferences for risk-taking and treatment effects in DMD. Real option value implies that an optimally informed, forward-looking patient could consider existing treatments and those that are in the pipeline in their current treatment decision-making [35]. Health economists indicated that real option value should probably not be prioritized as an element of value in a value assessment framework for DMD therapies because the likelihood of better future therapies would be based mainly on conjecture, not evidence. Payers may not consider real option value due to patient movement between healthcare plans or be willing to cover future high-cost therapies if patients have previously been treated with another drug. Patient advocates and clinicians discussed equity in the context of DMD and new DMD therapies, noting inequities in socioeconomic status, differences in health plans, and availability of health services across the Duchenne community, which affects access to care and resources. The health economists emphasized that equity can only be considered an element of value if disease incidence varies across different groups and the healthcare technology helps to reduce the disparity. Health inequity concerns have been incorporated into the economic evaluation of healthcare technologies using distributional CEA, which considers fairness in the distribution of costs and outcomes and the trade-offs that may occur between improving total health and equity and reducing unfair inequity in health across income levels [36]. Another approach would be to give a higher willingness-to-pay threshold to a treatment that reduces disparities.

Only the health economists identified insurance value as highly relevant to determining the value of DMD therapies, as this is a rare and severe disease. This element of value may apply to cell and gene therapies and could be estimated by determining what individuals would be willing to pay for an insurance premium offering them future access to therapies [37, 38]. GRACE analysis is relevant to insurance value. Treatments for diseases with a larger impact on quality of life are valued more; in traditional CEA, health improvements are valued the same, regardless of a patient’s baseline quality of life [39].

### Limitations

This initiative had several limitations. Some stakeholders were interviewed as a group. All three health economists were interviewed together, and two clinicians were interviewed together. These joint interviews could have introduced bias or inhibited conversation around an element of value or an associated topic of interest. Attempts to mitigate this included the use of the same format and slides for each interview and the same prompting questions. The preread material enabled all stakeholders to participate in the interviews with a good understanding of the ISPOR Value Flower, DMD disease severity, and the objective of the initiative, which facilitated the interview process. No payer stakeholder was interviewed. This was intentional, as payer opinions have been published in documents describing reimbursement decision-making; however, the payer perspective is an important part of the value discussion and may have provided additional insights. No manufacturer was interviewed. This was intentional to prevent any perception of bias around the inclusion of elements of value in a value assessment framework for new DMD therapies that benefit the manufacturer. No individuals with Duchenne were interviewed, but their perspectives were reflected by the patient advocates, some of whom were parents/caregivers.

### Future Implications

To thoroughly assess the value of new DMD therapies, there is the need to develop and implement a robust and comprehensive approach to data collection for each of the elements of value identified as impactful by the stakeholders included in this initiative. Data collection must involve engagement and collaboration with the Duchenne community [40], and may come from different sources, including clinical trials, observational studies, registries, or using modeling approaches. Data collection should begin with the elements of value that add the most benefit and can be most easily measured. DMD may be stratified into disease stages based on clinical characteristics and progression markers, including genetic testing, wheelchair usage, scoliosis treatment, or ventilation assistance [26]. Impacts should be associated with the various stages of DMD to capture the value of slowing or stopping the progression of disease.

## CONCLUSION

This initiative illustrates the work that must be done to ensure decision-makers properly assess value to make informed decisions about new DMD therapies and provide individuals with Duchenne access to the therapies they need. Thought leaders have proposed broadening and advancing approaches to value assessment frameworks for new healthcare technologies by including the economic and humanistic costs incurred by individuals with Duchenne, their caregivers, employers, and society. The patient advocates, clinicians, and health economists included in this initiative agreed that many of these costs, such as indirect out-of-pocket costs, lost productivity, and family spillovers, have important and compelling relevance for DMD. Some participants indicated that severity of disease, value of hope, and real option value are especially meaningful for individuals with disabilities and high unmet needs. Others noted the inclusion of equity, insurance value, and scientific spillovers could make improvements in population health. It became evident that a holistic approach that incorporates the perspectives of individuals with Duchenne, caregivers, clinicians, and heath economists is required to expand traditional value assessment frameworks, ensure accurate evaluations, and prevent restricted or delayed access to new therapies for individuals with Duchenne.

## Data Availability

Data sharing is not applicable to this article as no datasets were generated or analyzed during this study.

## ACKNOWLEDGMENTS

Funding for this paper was provided by Sarepta Therapeutics, Inc. Medical writing and editorial support were funded by Sarepta Therapeutics, Inc. The authors retained editorial control.

## AUTHOR CONTRIBUTIONS

**Conceptualization:** RF, PF, AK, KM, MP, DM, CL, LL, NLK, PBS, PJN, JS, RJW

**Methodology:** JK

**Validation:** RF, PF, AK, KM, MP, DM, CL, LL, PBS, PJN, JS, RJW

**Investigation:** RF, PF, AK, KM, MP, DM, CL, LL, NLK, PBS, JK, PJN, JS, RJW

**Data curation:** RF, PF, AK, KM, MP, DM, CL, LL, PBS, JK, PJN, JS, RJW

**Writing – original:** RF, PF, AK, KM, MP, DM, CL, LL, PBS, JK, PJN, JS, RJW

**Writing – review and editing:** RF, PF, AK, KM, MP, DM, CL, LL, NLK, PBS, JK, PJN, JS, RJW

**Project administration:** JK

## REFERENCES

[1] Bushby K, Finkel R, Birnkrant DJ, Case LE, Clemens PR, Cripe L, et al. Diagnosis and management of Duchenne muscular dystrophy, part 1: Diagnosis, and pharmacological and Psychosocial Management. Lancet Neurol. 2010;9(1):77–93. doi:10.1016/s1474-4422(09)70271-6

[2] Centers for Disease Control and Prevention-MMWR Weekly. https://www.cdc.gov/mmwr/preview/mmwrhtml/mm5840a1.htm#:~:text=Birth%20prevalence%20of%20DMD%20has,)%20male%20births%20(5). June 22, 2023.

[3] Ferizovic N, Summers J, de Zárate IB, Werner C, Jiang J, Landfeldt E, et al. Prognostic indicators of disease progression in Duchenne Muscular Dystrophy: A literature review and evidence synthesis. PLoS ONE. 2022;17(3). doi:10.1371/journal.pone.0265879

[4] Marden JR, Freimark J, Yao Z, Signorovitch J, Tian C, Wong BL. Real-world outcomes of long-term prednisone and deflazacort use in patients with Duchenne muscular dystrophy: Experience at a single, large care center. J Comp Eff Res. 2020;9(3):177–89. doi:10.2217/cer-2019-0170

[5] Birnkrant DJ, Bushby K, Bann CM, Alman BA, Apkon SD, Blackwell A, et al. DMD Care Considerations Working Group. Diagnosis and management of Duchenne muscular dystrophy, part 1: diagnosis, and neuromuscular, rehabilitation, endocrine, and gastrointestinal and nutritional management. Lancet Neurol. 2018;17(3):251–267. doi:10.1016/S1474-4422(18)30024-3.

[6] Duan D, Goemans N, Takeda S, Mercuri E, Aartsma-Rus A. Duchenne muscular dystrophy. Nat Rev Dis Primers. 2021;7(1):13. doi:10.1038/s41572-021-00248

[7] Filonova G, Aartsma-Rus A. Next steps for the optimization of exon therapy for Duchenne muscular dystrophy. Expert Opin Biol Ther. 2023;23(2):133–143. doi:10.1080/14712598.2023.2169070

[8] McDonald CM, Muntoni F, Penematsa V, Jiang J, Kristensen A, Bibbiani F, et al. Ataluren delays loss of ambulation and respiratory decline in nonsense mutation Duchenne muscular dystrophy patients. J Com Eff Res. 2022;11(3): 139–155. doi:10.2217/cer-2021-0196

[9] ELEVIDYS [package insert]. Cambridge (MA): Sarepta Therapeutics; 2023.

[10] Mandelblatt JS, Ramsey SD, Lieu TA, Phelps CE. Evaluating Frameworks That Provide Value Measures for Health Care Interventions. Value Health. 2017;20(2):185–192. doi:10.1016/j.jval.2016.11.013

[11] Brazier JE, Rowen D, Lloyd A, Karimi M. Future Directions in Valuing Benefits for Estimating QALYs: Is Time Up for the EQ-5D?. Value Health. 2019;22(1):62–68. https://doi.org/10.1016/j.jval.2018.12.001

[12] Garrison LP, Jr., Kamal-Bahl S, Towse A. Toward a Broader Concept of Value: Identifying and Defining Elements for an Expanded Cost-Effectiveness Analysis. Value Health. 2017;20(2):213–216. doi:10.1016/j.jval.2016.12.005

[13] Burkholder R, Dougherty JS, Neves LA. ISPOR’s Initiative on US Value Assessment Frameworks: An Industry Perspective. Value Health. 2018;21(2):173–175. doi:10.1016/j.jval.2017.12.001

[14] Sanders GD, Neumann PJ, Basu A, Brock DW, Feeny D, Krahn M, et al. Recommendations for conduct, methodological practices, and reporting of cost-effectiveness analyses: Second Panel on Cost-Effectiveness in Health and Medicine. JAMA. 2016;316(10):1093–1103. doi: 10.1001/jama.2016.12195

[15] Lakdawalla DN, Doshi JA, Garrison LP Jr, Phelps CE, Basu A, Danzon PM. Defining Elements of Value in Health Care-A Health Economics Approach: An ISPOR Special Task Force Report [3]. Value Health. 2018;21(2):131–139. doi:10.1016/j.jval.2017.12.007

[16] Neumann PJ, Garrison LP, Willke RJ. The History and Future of the “ISPOR Value Flower”: Addressing Limitations of Conventional Cost-Effectiveness Analysis. Value Health. 2022;25(4):558–565. doi:10.1016/j.jval.2022.01.010

[17] Garrison LP Jr, Lo AW, Finkel RS, Deverka PA. A review of economic issues for gene-targeted therapies: Value, affordability, and access. Am J Med Genet C Semin Med Genet. 2023;193(1):64–76. doi: 10.1002/ajmg.c.32037

[18] Yang G, Cintina I, Pariser A, Oehrlein E, Sullivan J, Kennedy A. The national economic burden of rare disease in the United States in 2019. Orphanet J Rare Dis. 2022;17(1). doi:10.1186/s13023-022-02299-5

[19] Leech AA, Lin PJ, D’Cruz B, Parsons SK, Lavelle TA. Family Spillover Effects: Are Economic Evaluations Misrepresenting the Value of Healthcare Interventions to Society? Appl Health Econ Health Policy. 2023 Jan;21(1):5–10. doi: 10.1007/s40258-022-00755-8. Epub 2022 Aug 23. PMID: 35997896; PMCID: PMC9839569.

[20] Han S, Xu H, Zheng J, Sun J, et al. Population-wide Duchenne Muscular Dystrophy carrier detection by CK and molecular testing. BioMed Research International. Article ID 8396429, 12 pages https://doi.org/10.1155/2020/8396429.

[21] Crossnohere NL, Fischer R, Lloyd A, Prosser LA, Bridges JFP. Assessing the Appropriateness of the EQ-5D for Duchenne Muscular Dystrophy: A Patient-Centered Study. Med Decis Making. 2021;41(2):209–221. doi: 10.1177/0272989X20978390

[22] Powell PA, Carlton J, Rowen D, Brazier J, Facey K, Bayley K, et al. Measuring What Matters: Little Evidence Supporting the Content Validity of EQ-5D in People with Duchenne Muscular Dystrophy and Their Caregivers. Med Decis Making. 2022;42(2):139–140. doi: 10.1177/0272989X211062237

[23] [Internet]. EveryLife Foundation for Rare Diseases; [cited 2023 Jun 6]. Available from: https://everylifefoundation.org/wp-content/uploads/2021/02/The_National_Economic_Burden_of_Rare_Disease_Study_Summary_Report_February_2021.pdf

[24] Social Security Act. Section 1182. [42 U.S.C. 1320e-1]

[25] Powell PA, Carlton J. A comprehensive qualitative framework for health-related quality of life in Duchenne Muscular Dystrophy. Qual Life Res. 2023;32(1):225–36. doi:10.1007/s11136-022-03240-w

[26] Iff J, Zhong Y, Gupta D, Paul X, Tuttle E, Henricson E, et al. Disease progression stages and burden in patients with Duchenne muscular dystrophy using administrative claims supplemented by electronic medical records. Adv Ther. 2022;39(6):2906–19. doi:10.1007/s12325-022-02117-1

[27] Labisa P, Andreozzi V, Mota M, Monteiro S, Alves R, Almeida J, et al. Cost of illness in patients with Duchenne muscular dystrophy in Portugal: The Coiduch Study. Pharmacoeconom Open. 2021;6(*2*):211–8. doi:10.1007/s41669-021-00303-5

[28] Soelaeman RH, Smith MG, Sahay K, Tilford JM, Goodenough D, Paramsothy P, et al. Labor market participation and productivity costs for female caregivers of minor male children with Duchenne and Becker muscular dystrophies. Muscle Nerve. 2021;64(6):717–25. doi:10.1002/mus.27429

[29] Perfetto EM, Oehrlein EM, Love TR, Schoch S, Kennedy A, Bright J. Patient-centered core impact sets: What they are and why we need them. Patient. 2022;15(6):619–27. doi:10.1007/s40271-022-00583-x

[30] NICE Health Technology Evaluations: The Manual [Internet]. NICE; [cited 2023 Jun 7]. Available from: https://www.nice.org.uk/process/pmg36/resources/nice-health-technology-evaluations-the-manual-pdf-72286779244741

[31] Peay HL, Fischer R, Mange B, Paquin RS, Smith EC, Sadosky A. Patients’ and caregivers’ maximum acceptable risk of death for nonLcurative gene therapy to treat Duchenne muscular dystrophy. Mol Genet Genomic Med. 2021;9(5). doi:10.1002/mgg3.1664

[32] Landrum Peay H, Fischer R, Tzeng JP, Hesterlee SE, Morris C, Strong Martin A, et al. Gene therapy as a potential therapeutic option for Duchenne Muscular Dystrophy: A qualitative preference study of patients and parents. PLoS ONE. 2019;14(5). doi:10.1371/journal.pone.0213649

[33] Peay HL, Hollin I, Fischer R, Bridges JFP. A community-engaged approach to quantifying caregiver preferences for the benefits and risks of emerging therapies for Duchenne muscular dystrophy. Clin Ther. 2014;36(5):624–37. doi:10.1016/j.clinthera.2014.04.011. Erratum in: Clin Ther. 2014 Aug 1;36(8):1313.

[34] Crossnohere NL, Fischer R, Vroom E, Furlong P, Bridges JF. A comparison of caregiver and patient preferences for treating Duchenne muscular dystrophy. Patient. 2022;15(5):577–88. doi:10.1007/s40271-022-00574-y

[35] Li M, Garrison L Jr, Lee W, Kowal S, Wong W, Veenstra D. A pragmatic guide to assessing real option value for medical technologies. Value Health. 2022;25(11):1878–84. doi.org/10.1016/j.jval.2022.05.014

[36] Asaria M, Griffin S, Cookson R. Distributional cost-effectiveness analysis. Med Decis Making. 2015;36(1):8–19. doi:10.1177/0272989X15583266

[37] Garrison LP, Jackson T, Paul D, Kenston M. Value-based pricing for emerging gene therapies: The economic case for a higher cost-effectiveness threshold. J Manag Care Spec Pharm. 2019;25(7):793–9. doi:10.18553/jmcp.2019.18378

[38] Drummond M, Ciani O, Fornaro G, Jommi C, Dietrich ES, Espin J, et al. How are Health Technology Assessment Bodies responding to the assessment challenges posed by cell and gene therapy? BMC Health Serv Rese. 2023;23(1). doi:10.1186/s12913-023-09494-5

[39] Lakdawalla DN, Phelps CE. Health Technology Assessment with diminishing returns to health: The generalized risk-adjusted cost-effectiveness (GRACE) approach. Value Health. 2021;24(2):244–9. doi:10.1016/j.jval.2020.10.003

[40] van Lin N, Paliouras G, Vroom E, ‘t Hoen PAC, Roos M. How patient organizations can drive fair data efforts to facilitate research and health care: A report of the virtual second international meeting on duchenne data sharing, March 3, 2021. Journal of Neuromuscul Dis. 2021;8(6):1097–108. doi:10.3233/jnd-210721

