## Supplementary material for "Healthcare Stakeholder Perspectives on a Value Assessment Approach for Duchenne Muscular Dystrophy Therapies": Table S1

### **Table S1**: Elements of Value [12-14]

| **Value Element** | **Definition** |
| --- | --- |
| **Quality-Adjusted Life Year (QALYs) Gained** | The QALY is the fraction of a perfectly healthy life-year that remains after accounting for the damaging effects of an illness or condition. |
| **Net Costs** | The intervention costs minus averted medical and productivity costs. |
| **Productivity** | Measuring and valuing productivity gains and losses due to healthcare technologies. |
| **Family Spillovers** | The impact of illness extends beyond the patient to unpaid caregivers and other family members who commonly incur out-of-pocket and time costs, lost productivity, and decreased HRQoL caring for and caring about a sick family member. |
| **Value of Knowing** | The value given to a healthcare technology that improves the certainty of outcomes or appropriate use of therapies. |
| **Insurance Value: Financial and Health** | The value of physical and financial risk protection of a new healthcare technology, whereby a new technology reduces the physical risk of getting or staying sick, offering “physical risk protection,” and the “financial risk protection” of greater options for consumers due to medical care, expanding the possibility of insuring against illness. |
| **Fear of Contagion and Disease** | This is most commonly associated with infectious diseases, and it assesses the benefits of a healthcare technology that extends beyond the treated patient. |
| **Severity of Disease** | Individuals place a greater weight (for themselves) on improvements in health from more severe health states than on equivalent improvements from less severe states. |
| **Value of Hope** | General population and /or patient perceived trade-offs for a chance of cure or extended survival. |
| **Real Option Value** | Real option value is generated when a healthcare technology that extends life creates opportunities for the patient to benefit from other future advances in medicine. |
| **Equity** | The value of healthcare technologies that address equity across populations, including healthy versus sick. |
| **Scientific Spillovers** | The value of the impact a new healthcare technology will have on the development of other healthcare technologies in the future. |

Note: The original ISPOR Value Flower included ‘adherence-improving factors’ as an element of value. In current developments, this element of value has been replaced with ‘family spillovers.’
