## Supplementary figures and images for "Healthcare Stakeholder Perspectives on a Value Assessment Approach for Duchenne Muscular Dystrophy Therapies"

### Figure S1

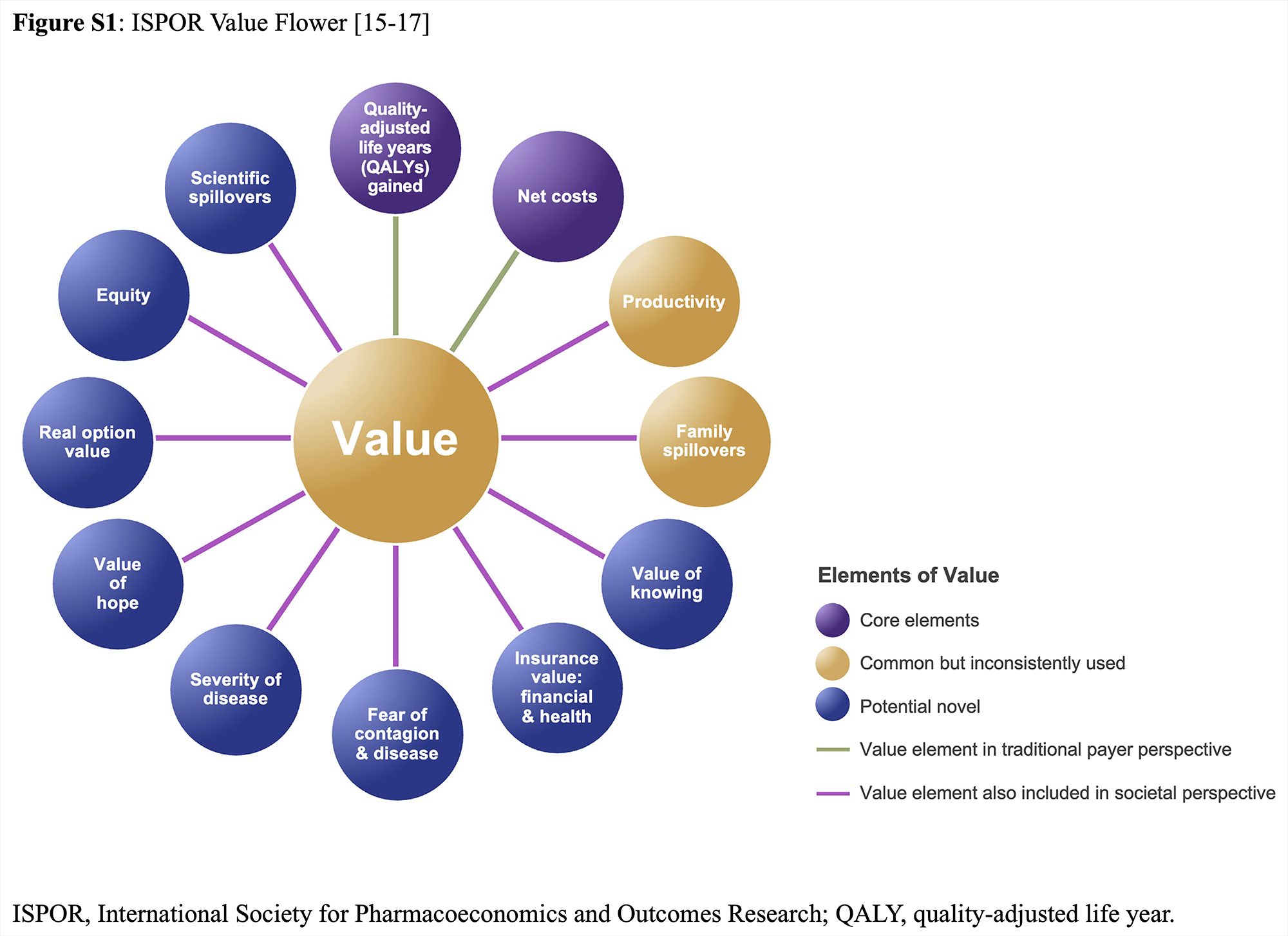
